# Knowledge, attitudes, practices of primary and middle school students at the outbreak of COVID-19 in Beijing: A cross-sectional online study

**DOI:** 10.1101/2020.06.29.20138628

**Authors:** Fuyuan Wen, Yi Meng, Han Cao, Juan Xia, Hui Li, Han Qi, Kai Meng, Ling Zhang

## Abstract

**Purpose:** This study investigated the KAP towards COVID-19 and their influencing factors among primary and middle school students during the self-quarantine period in Beijing.

**Methods:** This was a cross-sectional study among students from 18 primary and middle schools in Beijing during March 2020. Stratified cluster sampling was conducted. Demographic and KAP-related COVID-19 information was collected through an online questionnaire. The influencing factors were analyzed by multivariable logistic regression.

**Results:** A total of 7,377 students were included. The overall correct rate for COVID-19 knowledge was 74.1%, while only 31.5% and 40.5% could identify the high-risk places of cross-infection and warning body temperature. Although 94.5% of respondents believed the epidemic could be controlled, over 50% expressed various concerns about the epidemic. The compliance rates for basic preventing behaviors were all over 80%, while those for “rational and effective ventilation” (39.2%) and “dinning separately” (38.6%) were low. The KAP levels were significantly differed according to various school categories of students. The COVID-19 knowledge (OR= 3.309, 95% CI: 2.921, 3.748) and attitude (OR=1.145, 95% CI: 1.003, 1.308) were associated with preventive practices. Besides, female, urban students, those with a healthy lifestyle, and those with the willingness to engage in healthcare tended to have better preventive practices.

**Conclusion:** Most students in Beijing hold a high level of knowledge, optimistic attitudes and have appropriate practices towards COVID-19. However, targeted interventions are still necessary, especially for students with high-risk characteristics.

**Implications and contributions:** The performance and the potential factors of COVID-19-related knowledge, attitudes and practices (KAP) among students in primary and middle schools is still unclear.

This study investigates the characteristics and the level of KAP among students. The results of the study may contribute to the targeted education and interventions for students.

## 1. Introduction

An emerging respiratory disease, coronavirus disease 2019 (COVID-19), was detected in Wuhan and spread rapidly all over China [1]. On 11 March 2020, the World Health Organization (WHO) declared COVID-19 as a pandemic considering its rapid and broad spread ability [2]. The global cumulative number of confirmed cases was 2,719,897, and the death toll reached 187,705 as of April 25, 2020 [3]. Chinese government categorized COVID-19 as a Category B infectious disease and taking preventive and control measures for a Category A infectious disease [4]. From January 24, Beijing and other provinces and cities nationwide launched the first-level response mechanism for major public health emergencies, extending the winter vacation for all students so they could self-quarantine at home.

Currently, the COVID-19 epidemic has been well controlled in China, and primary and middle schools in various provinces and cities have been preparing for the start of school. School is considered as a high-risk for cross-infection place due to the concentrations of vulnerable groups and high social activity. In addition, primary and middle school students are a large and special group heavily affected by the epidemic in various aspects such as learning, education, life, etc., thereby being the focus of attention and protection during the epidemic. However, the vaccine for COVID-19 is still in the development phase [5], and the non-medical means (i.e., wash hands frequently, maintain a safe social distance and wearing the mask) is still the only option to individual protection. Previous studies have investigated the awareness, psychological and behavioral conditions among ordinary residents [6-15], healthcare professionals [16-21], and medical students [22-24] during the COVID-19 epidemic. A study about knowledge, attitudes, and practices (KAP) by Zhong et al. found that residents with a high level of knowledge on and positive attitudes toward COVID-19 tend to have better preventive behaviors and behavioral compliance [6]. However, the KAP of primary and middle school students towards COVID-19 remains sparse.

In this study, we investigated the KAP and related influencing factors towards COVID-19 among primary and middle school students through an online questionnaire survey during the self-quarantine period in Beijing. This study will provide scientific evidence and recommendations for educational institutions to carry out targeted health education measures, prevent the spread of the virus in schools, and ensure a smooth start to school.

## 2. Methods

### 2.1 Study participants

From March 2 to 13, 2020, we conducted a questionnaire survey with primary and middle school students in self-quarantine in Beijing during the COVID-19 epidemic. According to the principle of stratified cluster sampling, 2 districts were sampled from Beijing in which students from 5 primary schools, 6 junior high schools, 4 high schools, and 3 vocational high schools were selected as the subjects. The number of respondents for each grade was not less than 100. Students who were recruited through their school or the Internet and agreed to voluntary participate were asked to fill out the questionnaire online powered by www.wjx.cn, which is an online crowdsourcing platform in mainland China.

This study was approved by the Medical Ethical Committee in Beijing Anding Hospital of the Capital Medical University, China. Informed consent was confirmed by participants before the online survey, and all the participants could quit the investigation at any time.

### 2.2 Data collection

With reference to the latest research results and guidelines for COVID-19 that have been officially published at home and abroad and based on the design scheme of similar questionnaires [13, 25], we designed a questionnaire for primary and middle school students that was used after being reviewed and modified by experts. The items in the questionnaire included general demographic characteristics (household registration, gender, years in school, and place of residence at the time of the epidemic) as well as COVID-19-related knowledge (pathogen knowledge, epidemiological knowledge, and awareness of prevention and control measures), attitude (degree of concern and seriousness of the epidemic situation and confidence in defeating the epidemic), and preventive actions (actual practices).

The questionnaire included single-choice questions and multiple-choice questions. Single-choice questions only had 1 correct answer, with 1 point for each question; multiple-choice questions had multiple correct answers, with 2 points for each question when answered correctly, 0 points when answered with “I don’t know” or incorrectly, 2 points when no wrong choice was made and the number of correct choices for the item exceeded 60% of the correct choices for the item, and 1 point when the number of correct choices for the item was less than 60% of the correct choices for the item. There was a total of 23 items for COVID-19-related knowledge, including 13 single-choice questions and 10 multiple-choice questions, for a total score of 33 points.

According to the overall level of students’ answers, a score of 23-33 points was deemed as a high level of knowledge regarding COVID-19, and a score of 0-22 was deemed a low level of knowledge. For COVID-19-related attitude items, the responses were scored as follows: <2 point, negative attitude and ≥ 2 points, positive attitude. For COVID-19-related behavior questions, 11 preventive behaviors were included, each of which had a scored value of 1 point, for a total of 11 points; a total score ≥ 8 points indicated good practices, and a total score < 8 points indicated inappropriate practices.

### 2.3 Quality control

To ensure quality, 1 respondent from each unique IP address could respond to 1 questionnaire. The time to complete each questionnaire was automatically monitored through the network platform; respondents who completed the questionnaire in less than 200 seconds were excluded. Questionnaires answered by students not from primary and middle schools in Beijing, by students below the fourth grade, or by students who showed obvious inconsistency between grade and age were also excluded.

### 2.4 Statistical analysis

We characterized the distributions for all variates as the mean and standard deviation (SD) for continuous variables, and the count and percentage for categorical variables. Difference in knowledge scores and attitudes and practices for students with different characteristics were determined using the chi-square test or analysis of variance (ANOVA). Multiple logistic analysis was performed to analyze the associations of various characteristics with the KAP towards COVID-19. Odds ratios (ORs) and their 95% confidence intervals (CIs) were used to quantify the associations between variables and KAP. Additionally, we conducted a multivariate linear regression to calculate the contribution of each item of knowledge part toward the overall knowledge score. All statistical analyses were performed using IBM SPSS Statistics 25.0. Two-sided analysis was adopted, and P < 0.05 was considered as statistically significant.

## 3. Results

### 3.1 Characteristics of the subjects

A total of 8,864 questionnaires were collected, of which 7,377 (83.2%) were valid. The average age was 14.1 years old (range from 9 to18), A total of 3,577 (48.5%) were male, 3,213 (43.6%) were in junior high school, 4,760 (64.5%) lived in urban areas.

### 3.2 knowledge regarding COVID-19

Table 1 summarized the correct rates for COVID-19 related knowledge in primary and middle school students. Totally, the correct rate for COVID-19 related knowledge was 74.1%, among which it is over 80% for epidemiology (K1-K3), etiology (K7-K8) and prevention (K10-K16) knowledge of COVID-19, while only 31.5% and 40.5% could identify the high-risk places of cross-infection (K23) and warning body temperature (K22). There was a significant difference for COVID-19-related knowledge between various school categories (Table 1, Figure 1A). The correct rates of most knowledge were the highest in primary school students, while being the lowest in vocational school students (P < 0.001).

**Table 1.**
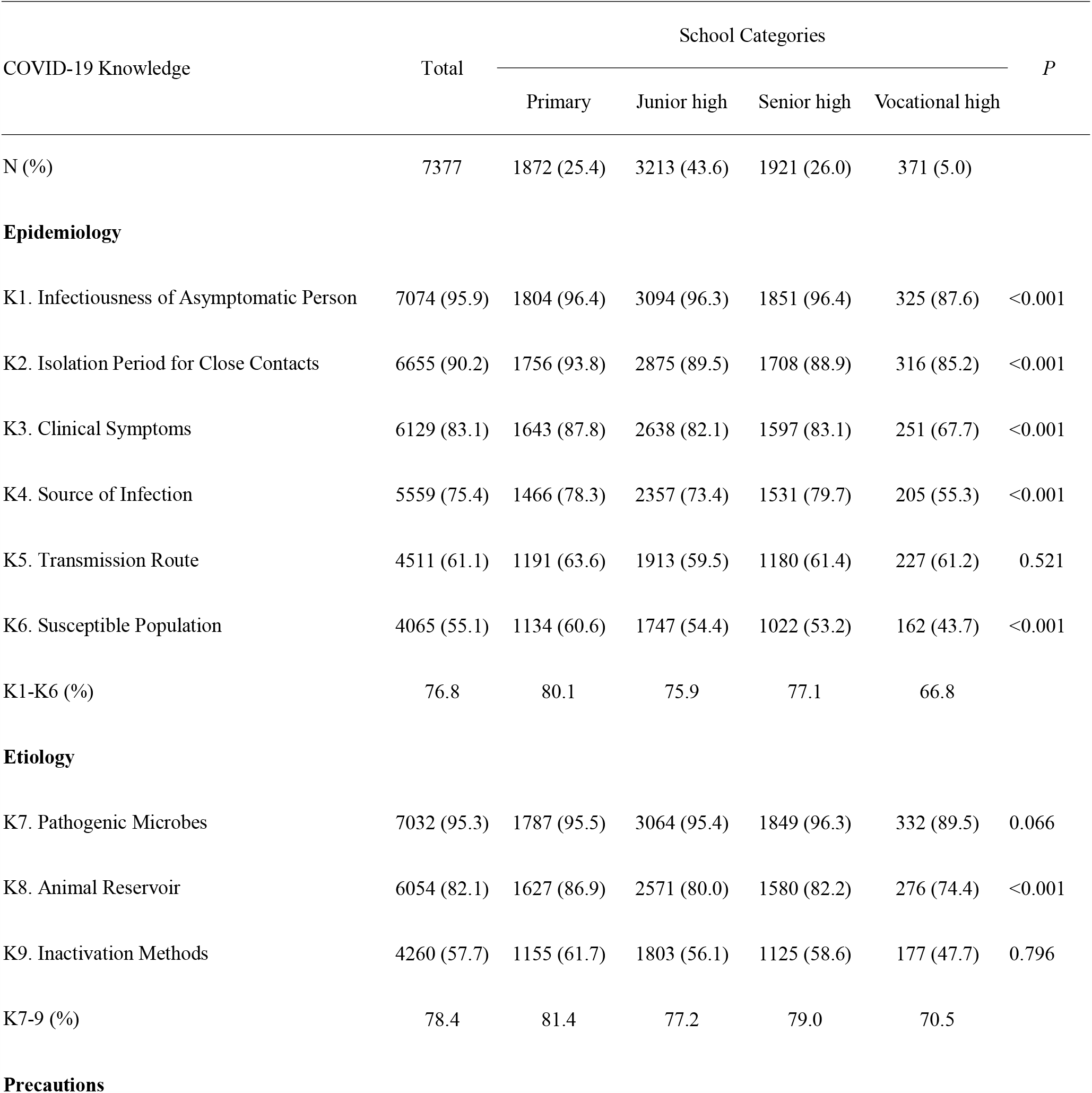
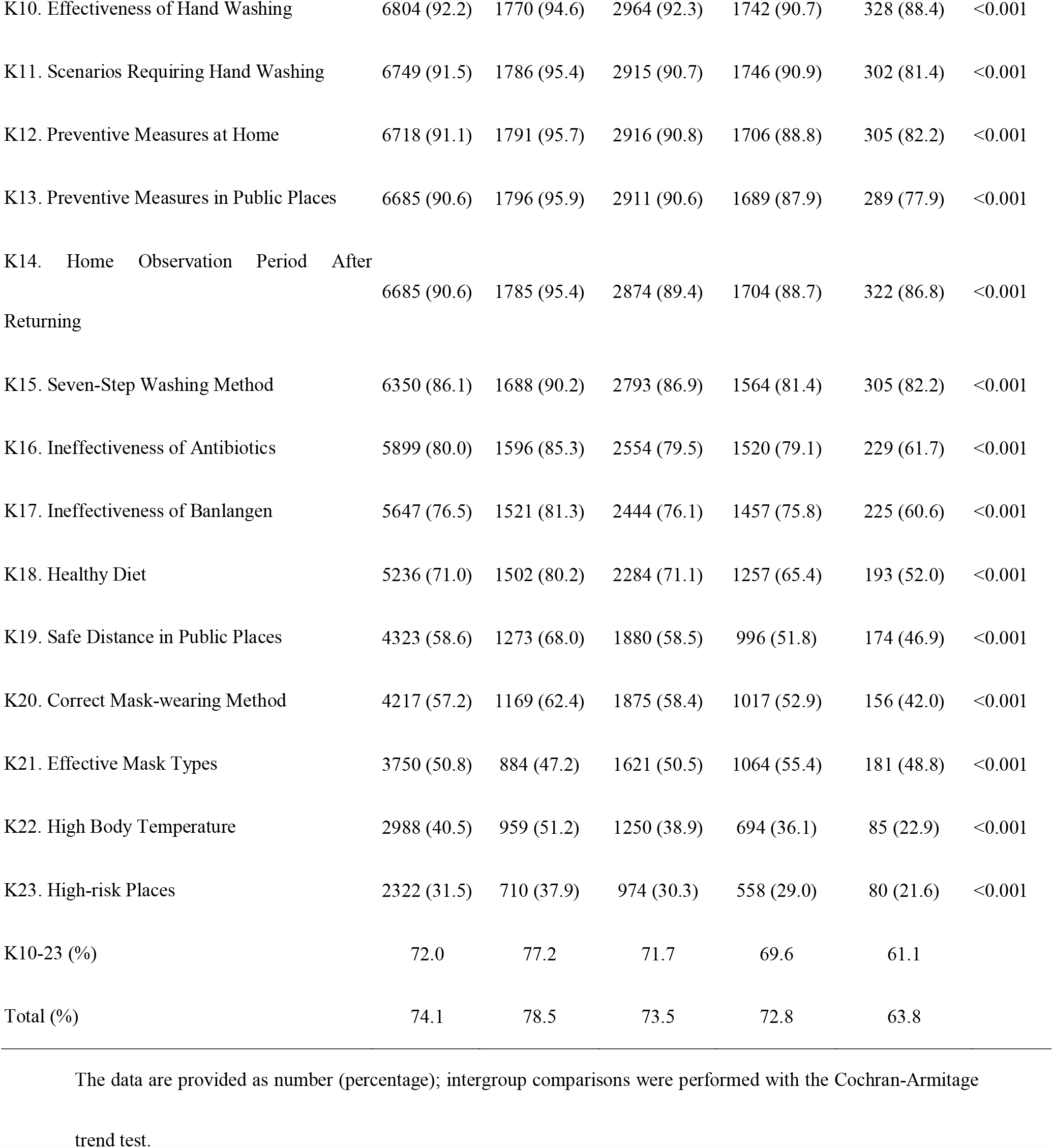
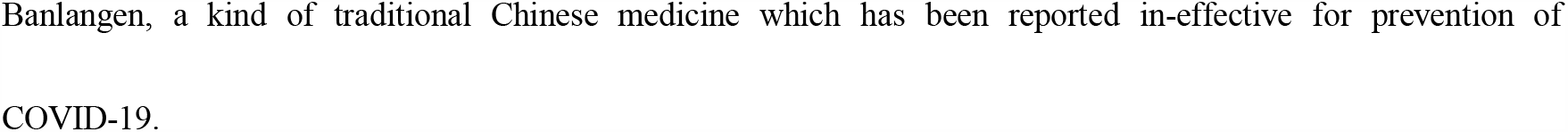
Correct rate for each item regarding COVID-19 knowledge by school categories (N, %)

**Figure 1.**
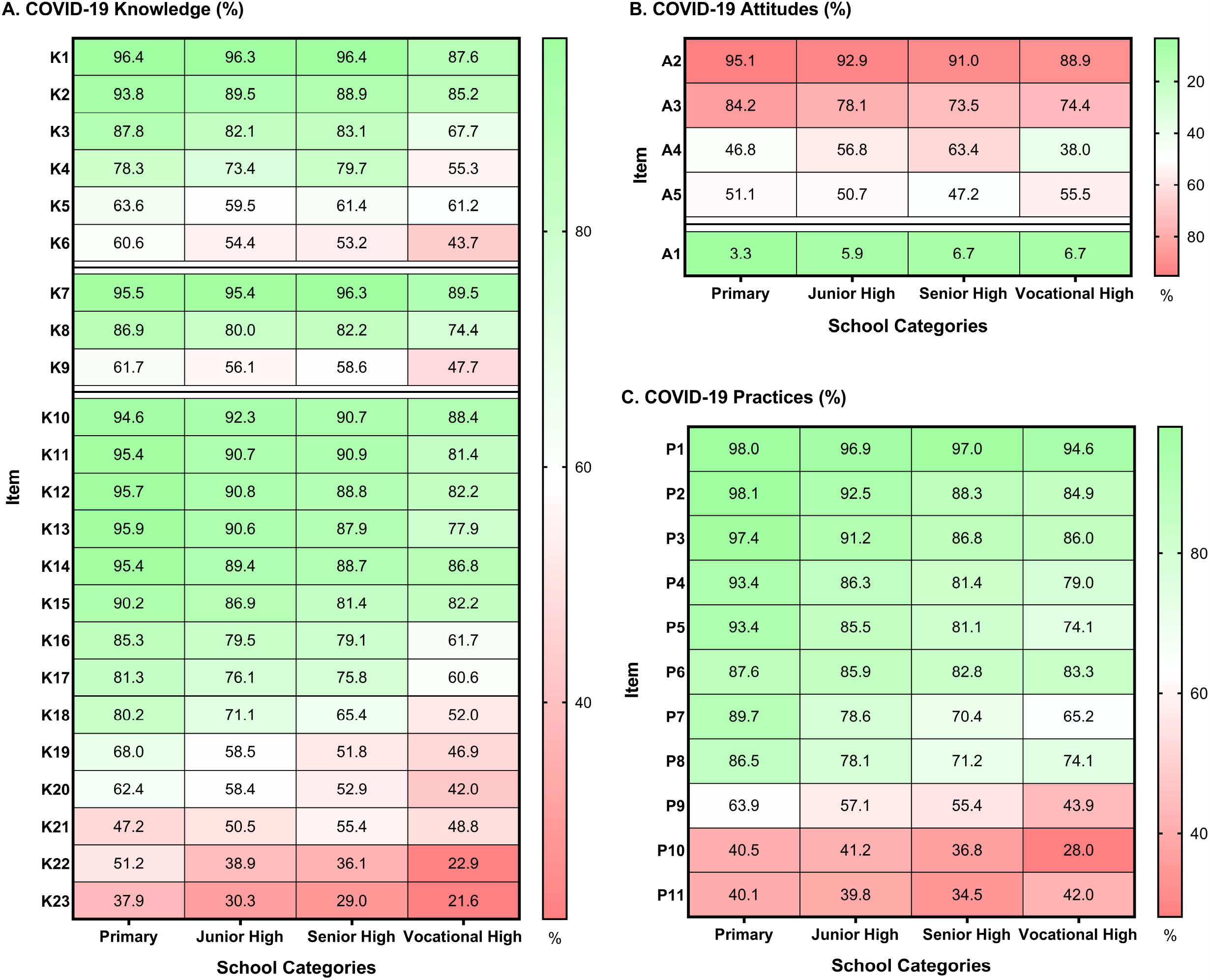
Correct rate for each item regarding COVID-19-related knowledge, attitudes and practices for students in different school categories. The results are shown as the rate for each item regarding COVID-19-related knowledge, attitudes and practices for different school categories; details for each item are available in Table 1–3. For each outcome, the color range of the cell, from red to green, represents worse to better. Students in different school categories share the same color scheme. COVID-19 knowledge, attitudes and practices are reported as proportions.

### 3.3 Attitude toward COVID-19

The attitude of the students toward COVID-19 was investigated through 5 questions, and the results showed that 94.5% of the students believed that “the epidemic will be controlled” (A1), 92.8% of the students believed that “the consequences of the infection will be very serious” (A2), 78.3% of the students were worried about being infected (A3), 55.0% of the students believed that “the epidemic will affect their academic performance” (A4), and 50.1% of the students felt nervous, scared and anxious about the epidemic (A5). The differences in A1-4 between students in different school categories were statistically significant (Table 2, Figure 1B).

**Table 2.**
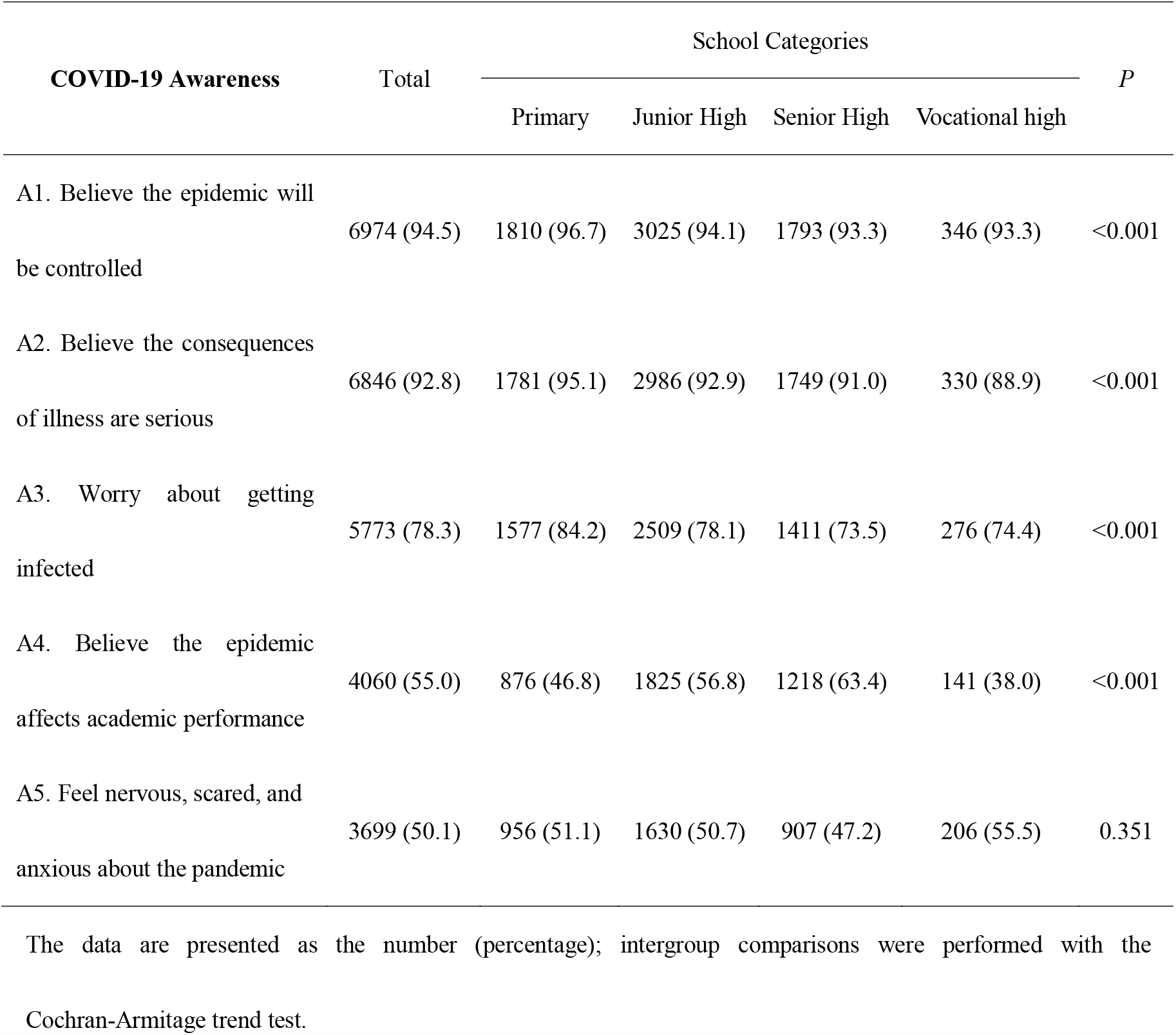
COVID-19 related attitudes by school categories (N, %)

| COVID-19 Awareness | Total | School Categories |  |  |  | *P* |
| --- | --- | --- | --- | --- | --- | --- |
|  |  | Primary | Junior High | Senior High | Vocational high |  |
| A1. Believe the epidemic will be controlled | 6974 (94.5) | 1810 (96.7) | 3025 (94.1) | 1793 (93.3) | 346 (93.3) | <0.001 |
| A2. Believe the consequences of illness are serious | 6846 (92.8) | 1781 (95.1) | 2986 (92.9) | 1749 (91.0) | 330 (88.9) | <0.001 |
| A3. Worry about getting infected | 5773 (78.3) | 1577 (84.2) | 2509 (78.1) | 1411 (73.5) | 276 (74.4) | <0.001 |
| A4. Believe the epidemic affects academic performance | 4060 (55.0) | 876 (46.8) | 1825 (56.8) | 1218 (63.4) | 141 (38.0) | <0.001 |
| A5. Feel nervous, scared, and anxious about the pandemic | 3699 (50.1) | 956 (51.1) | 1630 (50.7) | 907 (47.2) | 206 (55.5) | 0.351 |

The data are presented as the number (percentage); intergroup comparisons were performed with the Cochran-Armitage trend test.

### 3.4 Preventive behaviors during the COVID-19 epidemic

Table 3 showed the overall situation of the students regarding COVID-19-related preventive behaviors and differences between various school categories. The compliance rates for students regarding P1-6 were all above 80%, while “dining separately” (P11) were the lowest (38.6%). The compliance to effective preventive behaviors of primary school students was generally superior to that of students in high school (Table 3, Figure 1C).

**Table 3.**
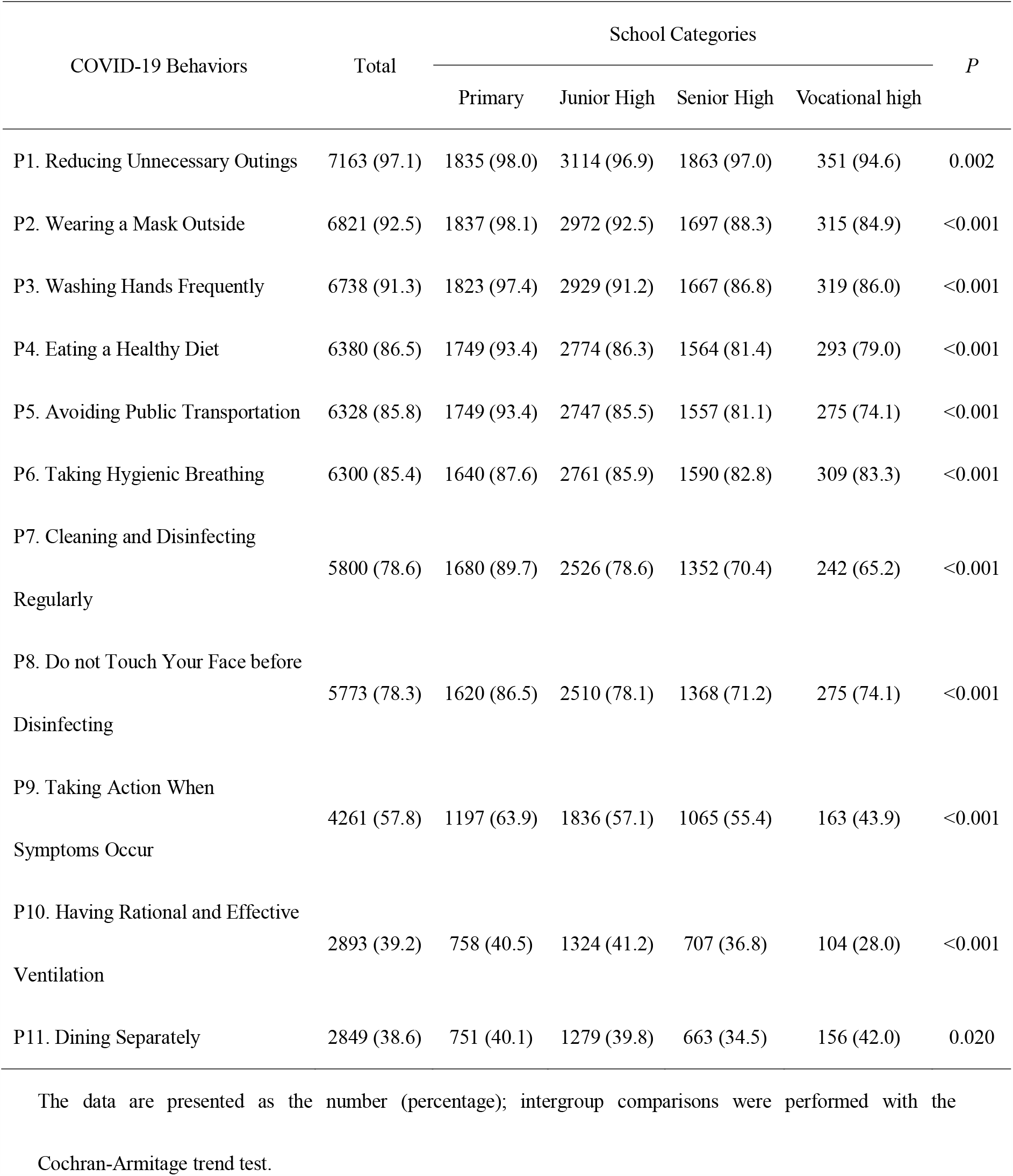
Compliance rate for each item regarding COVID-19-related practices by school categories (N,%)

### 3.5 Scores of KAP regarding COVID-19 according to different characteristics

The average scores of KAP for COVID-19 were 25.15 ± 3.86, 2.18 ± 0.90, and 8.31± 2.04, respectively (Table 4). The levels of KAP are significantly different in gender, school categories, residence, exercise duration, study duration, sleep duration, and willingness to become a healthcare professional in the future (P < 0.05).

**Table 4.** Scores of knowledge, attitudes, and practices toward COVID-19 according to different characteristics

| Variable |  | N (%) | Knowledge | *P* | Attitudes | *P* | Practices | *P* |
| --- | --- | --- | --- | --- | --- | --- | --- | --- |
| Total | - | 7377 (100.0) | 25.15 ± 3.86 |  | 2.18 ± 0.90 |  | 8.31 ± 2.04 |  |
| Age (years) | 9-12 | 1829 (24.8) | 26.11 ± 3.46 | <0.001 | 2.19 ± 0.91 | 0.277 | 8.88 ± 1.47 | <0.001 |
|  | 13-15 | 3190 (43.2) | 24.97 ± 3.99 |  | 2.16 ± 0.89 |  | 8.35 ± 2.06 |  |
|  | 16-18 | 2358 (32.0) | 24.63 ± 3.84 |  | 2.20 ± 0.91 |  | 7.81 ± 2.26 |  |
| Gender | Male | 3577 (48.5) | 24.75 ± 4.07 | <0.001 | 2.13 ± 0.89 | <0.001 | 8.08 ± 2.27 | <0.001 |
|  | Female | 3800 (51.5) | 25.52 ± 3.61 |  | 2.24 ± 0.91 |  | 8.53 ± 1.77 |  |
| School Categories | Primary | 1872 (25.4) | 26.11 ± 3.48 | <0.001 | 2.19 ± 0.90 | <0.001 | 8.89 ± 1.49 | <0.001 |
|  | Junior high | 3213 (43.6) | 24.95 ± 3.98 |  | 2.16 ± 0.89 |  | 8.33 ± 2.06 |  |
|  | Senior high | 1921 (26.0) | 24.99 ± 3.72 |  | 2.18 ± 0.92 |  | 7.86 ± 2.24 |  |
|  | Vocational high | 371 (5.0) | 22.78 ± 3.96 |  | 2.36 ± 0.90 |  | 7.55 ± 2.36 |  |
| Residence | Urban | 4760 (64.5) | 25.58 ± 3.73 | <0.001 | 2.21 ± 0.91 | <0.001 | 8.51 ± 1.89 | <0.001 |
|  | Rural | 2617 (35.5) | 24.35 ± 3.95 |  | 2.13 ± 0.89 |  | 7.94 ± 2.24 |  |
| Learning Duration | 1-2 h/d | 441 (6.0) | 24.81 ± 4.09 | <0.001 | 2.16 ± 0.96 | 0.330 | 8.13 ± 2.23 | 0.026 |
|  | 3-4 h/d | 1479 (20.0) | 25.19 ± 3.79 |  | 2.22 ± 0.91 |  | 8.39 ± 1.94 |  |
|  | 5-7 h/d | 2815 (38.2) | 25.30 ± 3.85 |  | 2.18 ± 0.89 |  | 8.28 ± 2.05 |  |
|  | 8-12 h/d | 1915 (26.0) | 25.22 ± 3.73 |  | 2.16 ± 0.88 |  | 8.38 ± 1.94 |  |
|  | >12 h/d | 727 (9.9) | 24.48 ± 4.16 |  | 2.21 ± 0.94 |  | 8.19 ± 2.29 |  |
| Exercise Duration | Never | 165 (2.2) | 22.95 ± 4.48 | <0.001 | 2.04 ± 0.91 | <0.001 | 6.79 ± 2.57 | <0.001 |
|  | <30 min/d | 2225 (30.2) | 25.03 ± 3.88 |  | 2.11 ± 0.92 |  | 7.95 ± 2.11 |  |
|  | 30-60 min/d | 3745 (50.8) | 25.34 ± 3.76 |  | 2.21 ± 0.88 |  | 8.50 ± 1.89 |  |
|  | >60 min/d | 1242 (16.8) | 25.06 ± 3.92 |  | 2.27 ± 0.92 |  | 8.60 ± 2.09 |  |
| Sleep Duration | <6 h/d | 256 (3.5) | 23.08 ± 4.69 | <0.001 | 2.11 ± 0.97 | 0.373 | 6.99 ± 2.67 | <0.001 |
|  | 6-8 h/d | 3249 (44.0) | 24.98 ± 3.77 |  | 2.17 ± 0.90 |  | 8.11 ± 2.08 |  |
|  | 8-10 h/d | 3451 (46.8) | 25.53 ± 3.78 |  | 2.20 ± 0.89 |  | 8.56 ± 1.89 |  |
|  | >10 h/d | 421 (5.7) | 24.55 ± 4.09 |  | 2.18 ± 0.91 |  | 8.66 ± 1.94 |  |
| Healthcare Occupation | Willing | 5397 (73.2) | 25.29 ± 3.79 | <0.001 | 2.20 ± 0.89 | 0.006 | 8.48 ± 1.97 | <0.001 |
|  | Unwilling | 1980 (26.8) | 24.75 ± 4.02 |  | 2.14 ± 0.93 |  | 7.84 ± 2.14 |  |

The data are presented as N (%) or Mean ± SD. One-way ANOVA was used to test for differences among continuous variables.

### 3.6 Multiple logistic regression analyses of KAP related factors

As shown in Table 5, about 77.4% of the students showed a high level of knowledge (with a score ≥ 23), 77.1% of the students had a positive attitude (with a score ≥ 2), and 75.6% of students had favorable preventive practices regarding COVID-19 (with a score ≥ 8), respectively. The results of multivariate logistic analyses showed that the level of COVID-19 knowledge was associated with gender, school categories, residence, willingness to become a healthcare professional, and lifestyle of exercise duration, sleep duration, learning duration; The level of attitude toward COVID-19 was associated with gender, residence, willingness to become a healthcare profession, exercise duration and level of COVID-19 knowledge (OR= 1.179, 95% CI: 1.035, 1.344). The favorable preventive behaviors were performed better among students with a higher level of COVID-19 knowledge (OR= 3.309, 95% CI: 2.921, 3.748) and positive attitude (OR=1.145, 95% CI: 1.003, 1.308), female students (OR=1.464, 95% CI: 1.303, 1.644), participants who were lived in urban out of Beijing during the COVID-19 outbreak (OR=1.372, 95% CI:1.217, 1.546), and students who willing to engage in the healthcare profession (OR=1.618, 95% CI:1.431, 1.830). Compared to students who did not exercise, better preventive behaviors were performed among those exercised less than 30 minutes/day (OR=1.445, 95% CI:1.016, 2.055), 30-60 minutes/day (OR=2.179, 95%CI: 1.531-3.099) and more than 60 minutes/day (OR=2.413, 95%CI: 1.658-3.513). Compared to primary school students, poor preventive behaviors were performed among those in junior high school (OR=0.622, 95% CI: 0.517, 0.748), senior high school (OR=0.437, 95% CI: 0.356, 0.537) and vocational high school (OR=0.556, 95% CI: 0.422, 0.731).

**Table 5.**
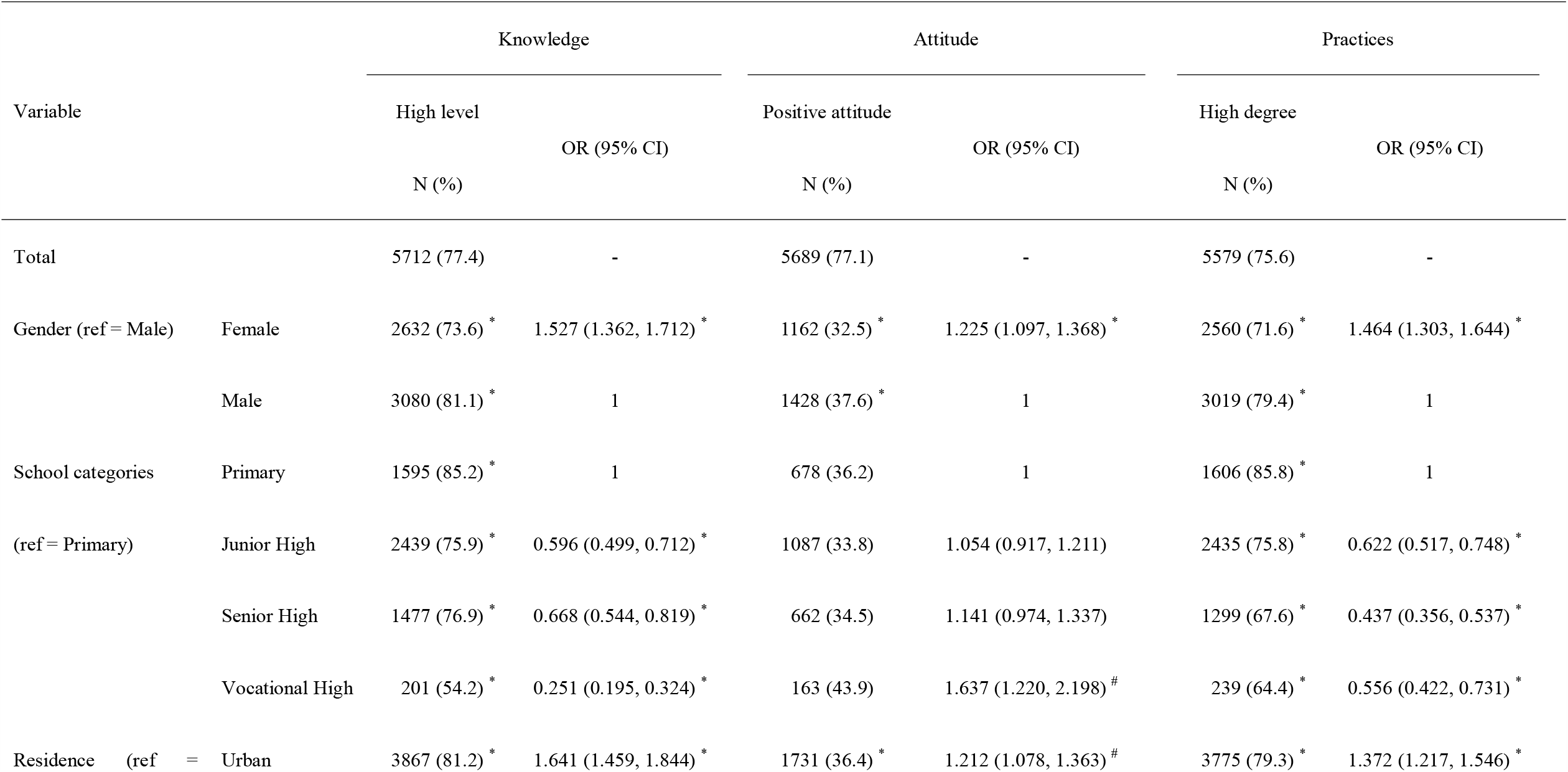
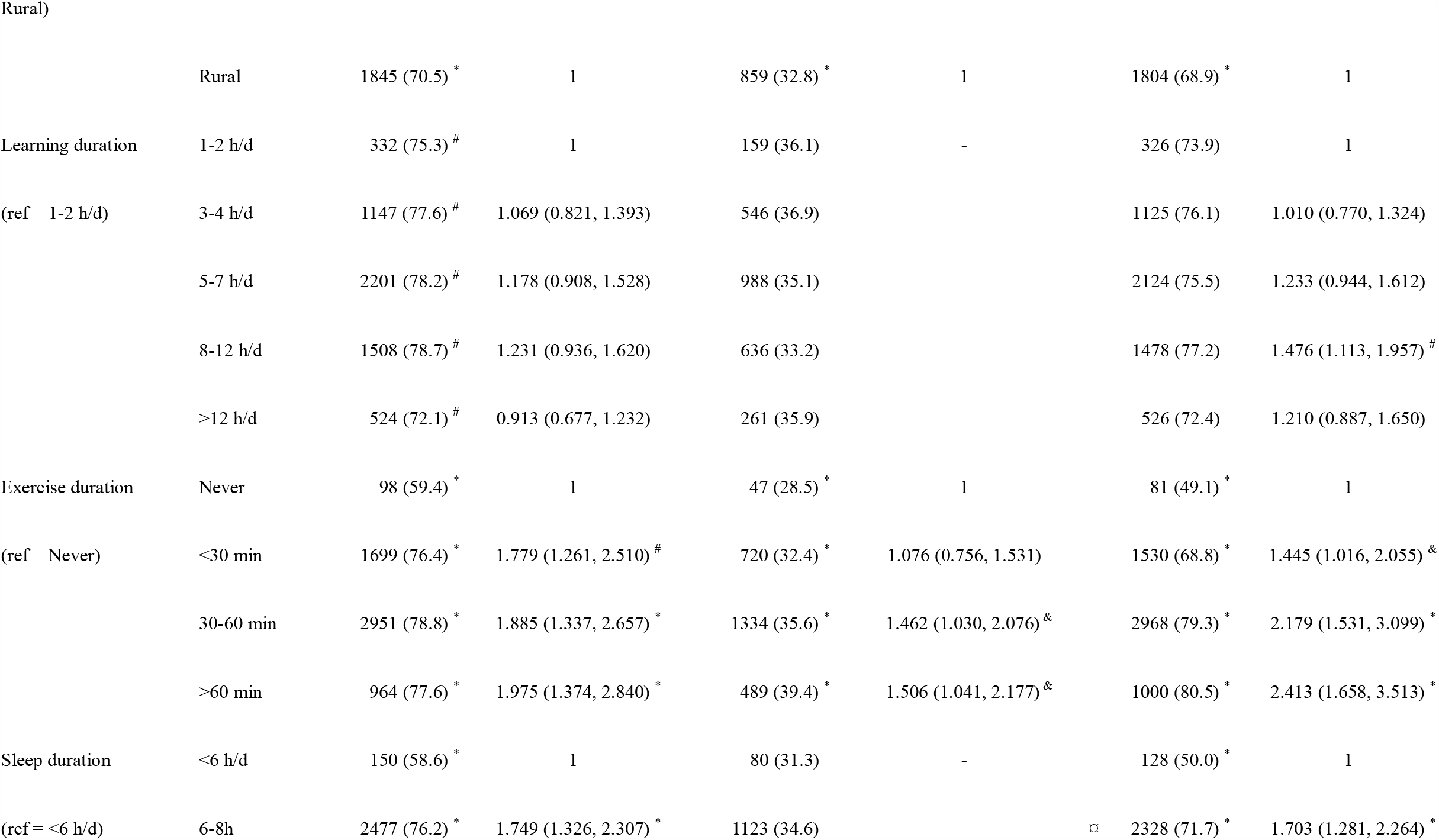
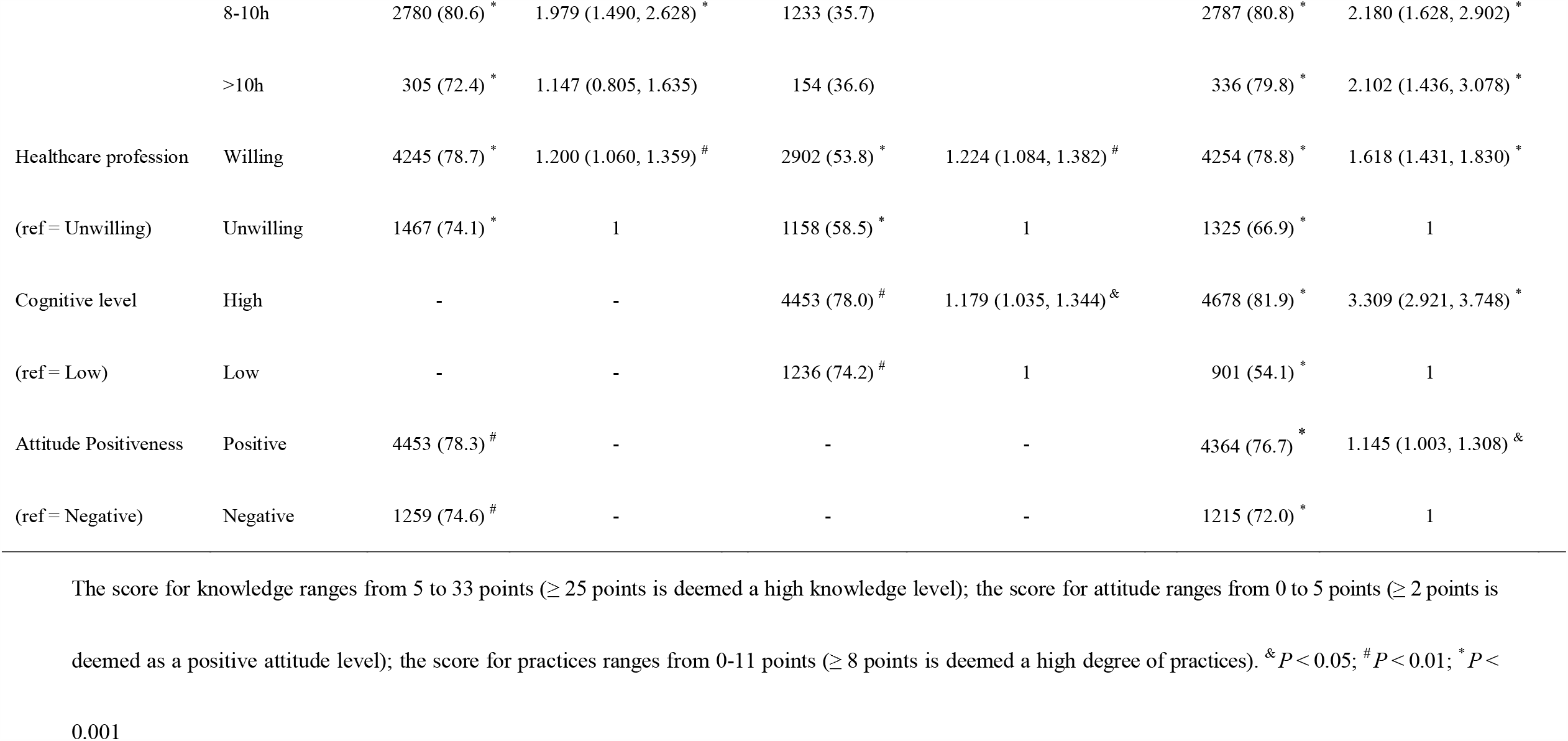
Multiple logistic regression analysis of factors significantly associated with knowledge, attitudes and practices regarding COVID-19

## 4. Discussion

To our knowledge, this is the first study examining the KAP and their influencing factors towards COVID-19 among primary and middle school students in Beijing. Our results show that the most of students are knowledgeable about COVID-19 and have optimistic attitudes and appropriate practices. In addition, the score of knowledge and attitudes towards COVID-19 were positively associated with preventive practices. Female, urban students, those with a healthy lifestyle, and those with the willingness to engage in healthcare tended to have better preventive behaviors.

In this study, the overall correct rate for students on knowledge was 74.1%, indicating that the majority of students acquired a certain understanding of COVID-19. We consider that this is primarily due to the effective education by the WHO and the Chinese government. Furthermore, students would actively learn knowledge of COVID-19 from various channels of information, such as CCTV and the official website of the National Health Commission of China since the serious situation of the epidemic and the overwhelming news reports on this public health emergency. However, the knowledge scope of students towards COVID-19 was not comprehensive. Although 95.9% of the students learned that asymptomatic carriers of the virus are contagious, only 61.1% and 55.1% knew the virus transmission routes and the susceptible populations. On the contrary, Geldsetzer and Chen et al. found that the residents had a high awareness level of the virus’s transmission routes in the United States, the United Kingdom, and Anhui province in China [7, 11]. Therefore, the knowledge about COVID-19 still needs to be strengthened through targeted education.

Besides, the results showed that primary and middle school students had a better level of knowledge regarding preventive measurement in washing hands. The WHO has proposed effective measures to prevent the spread of COVID-19 infection, including frequent hand washing, wearing a face mask when going out and keeping a safe social distance [26]. Hand washing is one of the most important and effective means of controlling the spread of the virus and preventing the development of infection at low cost [27]. However, the knowledge regarding other aspects such as maintaining proper social distance, wearing a face mask, monitoring body temperature closely, and identifying high-risk locations still needs to be improved.

In terms of attitudes, consistent with the results obtained from adults in China [6], more than 95% of the students believed that the epidemic would eventually be brought under control. However, more than half of the students were worried about being infected, and over 95% of the students believed that the consequences would be very serious once affected. Several studies were indicating that over concern about the COVID-19 epidemic was positively associated with the incidence of mental problems (e.g., anxiety and depression) [25, 28]. Although COVID-19 is highly contagious, most patients only manifest mild symptoms with a low overall crude mortality rate. And the death was tended to occur in susceptible populations, such as older people (more than 60 years) with underlying diseases [29]. Children and adolescents are also susceptible to COVID-19 infection but less likely to develop serious illnesses [30, 31]. Moreover, daily learning tasks have been completed through online teaching during the epidemic period. Therefore, psychological intervention implemented by the government and health authorities is recommended to avoid overloaded worrying about the epidemic.

The control of the COVID-19 epidemic in China is largely attributed to strict non-medical interventions, such as community disinfection, hand washing, isolation, movement restrictions and the use of masks [32]. In this study, we found that students were doing a good job in avoiding going out, wearing a face mask, washing hands, eating a healthy diet, avoiding public transportation, and covering the nose and mouth when sneezing. However, their compliance regarding separate dining and effective ventilation was still poor. Shared dishes is a common way of dining in China and is likely to facilitate the spread of the virus. Therefore, these weak behaviors need to be educated and corrected by schools and parents according to the guidelines given by the Ministry of Education of China [33].

Under the effective control of COVID-19 in China, the primary and middle schools are reopening recently. Protecting students fromCOVID-19 infection are calling for attention [34]. In this study, we found that the KAP level of COVID-19 in male students was significantly lower than those in females, which may be related to female students being more sensitive and concerned about changes in the external environment. Our study also revealed that students who lived in urban areas showed higher KAP levels than those who lived in rural areas, which is consistent with the observations in the previous studies [6, 8]. Furthermore, students in junior, senior and vocational high school showed lower levels of KAP than that in primary school students. School categories indirectly reflects the age of students in a certain way. A study conducted in young people in Hong Kong during the severe acute respiratory syndrome (SARS) epidemic period showed that the students at a younger age tend to show better preventive behaviors [35]. Vocational high school students are generally less interested in learning and thus poor at grasping knowledge. Due to heavy academic pressure, junior and senior high school students have little time to acquire COVID-19-related knowledge. Primary school students have lighter study loads, better learning compliance, and a stronger ability to accept new knowledge. Therefore, male, rural, and junior, senior and vocational high school students should call for more attention on health education. Interestingly, we found students with the willingness of becoming healthcare professionals have higher KAP levels regarding COVID-19. This may be because these students are more concerned about the progress of the COVID-19 epidemic. During the H1N1 epidemic, medical students outperformed the public in terms of knowledge, attitude and behavior [36]. Our study also revealed that those who exercise properly, get enough sleep, and study for an appropriate amount of time every day have higher levels of COVID-19-related KAP. Previous studies on influenza vaccination practices have also demonstrated that healthy lifestyles often lead to higher vaccination rates [37]. This finding suggests that a healthy lifestyle might be a potential factor of high-level KAP.

Additionally, we found that high knowledge levels and positive attitudes are influencing factors for the formation of better preventive practices, suggesting that developing good preventive behaviors requires knowledge about infectious diseases and that assuming a positive and correct attitude toward the COVID-19 epidemic is conducive to establishing good preventive behaviors. These observations are consistent with the results reported by Zhong et al. for Chinese residents and Zhou et al. for healthcare professionals in Wuhan [6, 19]. The government and education authorities can improve students’ psychological status and prevent bad behaviors by enhancing students’ knowledge level of COVID-19.

As the epidemic is well controlled in China, the resumption of classes for students is imminent. Understanding the KAP levels of primary and middle school students regarding COVID-19 can provide a basis for formulating relevant policies and targeted health education for the resumption of classes. Our results show that the majority of students in Beijing are knowledgeable about COVID-19, and have optimistic attitudes and appropriate practices. However, the knowledge scope of students towards COVID-19 was not comprehensive, resulting in over worrying about the epidemic. In addition, the health education for junior, senior and vocational high school and rural students are not enough. Therefore, governments and schools need to strengthen the education on students, help them maintain high prevention awareness, and adopt reasonable protective behaviors to avoid the COVID-19 after the school reopening.

The strengths of this study are as follows: (1) for the first time, a KAP survey regarding COVID-19 was conducted with a large sample of students in primary and middle schools; (2) the results of the stratified analysis of different school categories, i.e., primary school, junior, senior and vocational high school provides implications for improving health education that enables the resumption of school in a calculated manner; and (3) the survey covers a wide range of topics with multiple dimensions and thus provides evidence for a comprehensive understanding of the status and influencing factors of the KAP of primary and middle school students regarding COVID-19. However, this study also has certain limitations. First, the accuracy of the data collected through an online survey cannot be guaranteed. However, the home isolation period during the epidemic provided an opportunity to quickly obtain information through the convenience of an online survey, thus providing the time-sensitive research evidence for the timely adjustment of health education methods and the formulation of relevant epidemic prevention measures. Additionally, we adopted the cluster stratified sampling method while recruiting respondents through the publicity and mobilization efforts of the education system, which, to some extent, methodologically compensated for the inadequacy of the online survey. Second, this study only included children and adolescents who enrolled in school; therefore, the general characteristics of the population aged 9-18 years were incomplete. Nevertheless, the findings are of great significance for schools and other educational institutions. Lastly, this study was conducted in Beijing, the capital of China, during the COVID-19 outbreak. Because of the capital’s high level of education and its information delivery network, the overall performance of KAP we investigated might be higher than the mean level in China in general, which means that, in the future, multicenter studies involving large samples are needed nationwide to obtain more accurate data.

## 5. Conclusion

The most of students in Beijing are knowledgeable about COVID-19 and have optimistic attitudes and appropriate practices. The improvement of correct knowledge and positive attitudes may also lead to a better performance of preventive practices towards COVID-19. However, targeted interventions are still necessary, especially for male, rural and high school students.

## Data Availability

No additional data are available.

## Abbreviations

COVID-19: coronavirus disease 2019
KAP: Knowledge, attitudes and practices

## Acknowledgements

We thank all participants in this study.

## Data Sharing Statement

No additional data are available.

## Authors’ contribution

Fuyuan Wen and Yi Meng: database establishment, statistical analysis and writing of the manuscript.

Han Cao, Juan Xia, Hui Li: data collection and modification of the manuscript.

Kai Meng and Ling Zhang: study design, data collection, direction of statistical analysis and writing and modification of the manuscript.

